# The SARS-CoV-2 Alpha variant is associated with increased clinical severity of COVID-19 in Scotland: a genomics-based retrospective cohort analysis

**DOI:** 10.1101/2021.08.17.21260128

**Authors:** David J. Pascall, Elen Vink, Rachel Blacow, Naomi Bulteel, Alasdair Campbell, Robyn Campbell, Sarah Clifford, Chris Davis, Ana da Silva Filipe, Noha El Sakka, Ludmila Fjodorova, Ruth Forrest, Emily Goldstein, Rory Gunson, John Haughney, Matthew T.G. Holden, Patrick Honour, Joseph Hughes, Edward James, Tim Lewis, Samantha Lycett, Oscar MacLean, Martin McHugh, Guy Mollett, Yusuke Onishi, Ben Parcell, Surajit Ray, David L Robertson, Sharif Shabaan, James G. Shepherd, Katherine Smollett, Kate Templeton, Elizabeth Wastnedge, Craig Wilkie, Thomas Williams, The COVID-19 Genomics UK (COG-UK) consortium, Emma C. Thomson

## Abstract

**Objectives:** The SARS-CoV-2 Alpha variant was associated with increased transmission relative to other variants present at the time of its emergence and several studies have shown an association between Alpha variant infection and increased hospitalisation and 28-day mortality. However, none have addressed the impact on maximum severity of illness in the general population classified by the level of respiratory support required, or death. We aimed to do this.

**Methods:** In this retrospective multi-centre clinical cohort sub-study of the COG-UK consortium, 1475 samples from Scottish hospitalised and community cases collected between 1^st^ November 2020 and 30^th^ January 2021 were sequenced. We matched sequence data to clinical outcomes as the variant became dominant in Scotland and modelled the association between Alpha variant infection and severe disease using a 4-point scale of maximum severity by 28 days: 1. no respiratory support, 2. supplemental oxygen, 3. ventilation and 4. death.

**Results:** Our cumulative generalised linear mixed model analyses found evidence (cumulative odds ratio: 1.40, 95% CI: 1.02, 1.93) of a positive association between increased clinical severity and lineage (Alpha variant versus non-Alpha variant).

**Conclusions:** The Alpha variant was associated with more severe clinical disease in the Scottish population than co-circulating lineages.

## Introduction

The Alpha variant of SARS-CoV-2 (Pango lineage B.1.1.7) was first identified in the UK in September 2020 and has since been reported in 184 countries (1). It is defined by 21 genomic mutations or deletions, including 8 characteristic changes within the spike gene (Table S1) (2). These are associated with increased ACE-2 receptor binding affinity and innate and adaptive immune evasion (3-6) compared to preceding lineages. The Alpha variant, the first variant of concern (VOC), was estimated to be 50-100% more transmissible than others present at the time of its emergence (7), explaining the transient dominance of this lineage globally.

The presence of a spike gene deletion (Δ69-70) results in spike-gene target failure (SGTF) in real-time reverse transcriptase polymerase chain reaction (RT-PCR) diagnostic assays and provided a useful proxy for the presence of the Alpha variant for epidemiological analysis during this time period (2). Four large community analyses have shown a positive association between the presence of SGTF and 28-day mortality, with hazard ratios of 1.55 (CI 1.39-1.72), 1.64 (CI 1.32-2.04), 1.67 (CI 1.34-2.09) and 1.73 (CI 1.41-2.13) (8-10). Both SGTF (hazard ratios of 1.52 (CI 1.47-1.57), 1.62 (CI 1.48 - 1.78)) (11,12) and confirmed Alpha variant infection (hazard ratios of 1.34 (CI 1.07-1.66) and 1.61 (CI 1.28 - 2.03) (12,13) were associated with an increased risk of hospitalisation in community cases, and a smaller study of hospitalised patients found a greater risk of hypoxia at admission in those with confirmed Alpha variant infection (14). In contrast, other smaller analyses of hospitalised patients found no association between confirmed Alpha variant infection and increased clinical severity based on a variety of indices (15-17). Limited data are available on the full clinical course of disease with the Alpha variant in relation to other variants.

Understanding the clinical pattern of disease with new variants of concern is important for several reasons. Firstly, if a variant is more pathogenic than previous variants, this has implications for public health restrictions and the functioning of health care systems. Secondly, large numbers of low- and middle-income countries still have less than 50% of their populations having been vaccinated against SARS-CoV-2 (18). A better understanding of a variant with increased severity is important in modelling the impact of unmitigated infection in these settings. A clear understanding of the behaviour of the Alpha variant, which emerged as a dominant variant, is needed as a baseline to compare the clinical phenotype of later variants, such as the currently dominant Omicron sub-variant BA.5. Post-Alpha variants have been shown to be able to evade vaccine-induced immunity and therefore have the potential to spread even in immunised populations (19), so a historical understanding of severity remains important, as it seems unlikely that the SARS-CoV-2 infections will be brought under control in the near future.

We aimed to quantify the clinical features and rate of spread of Alpha variant infections in Scotland in a comprehensive national dataset. We used whole genome sequencing data to analyse patient presentations between 1^st^ November 2020 and 30^th^ January 2021 as the variant emerged in Scotland and used cumulative generalised additive models to compare 28-day maximum clinical severity for the Alpha variant against other lineages over the same period.

## Materials and Methods

### Sequencing

sequencing was performed as part of the COG-UK consortium using amplicon-based next generation sequencing (20,21).

### Bioinformatics

sequence alignment, lineage assignment and tree generation were performed using the COG-UK data pipeline (https://github.com/COG-UK/datapipe) and phylogenetic pipeline (https://github.com/cov-ert/phylopipe) with pangolin lineage assignment (https://github.com/cov-lineages/pangolin) (22). Lineage assignments were performed on 18/03/2021 and phylogenetic analysis was performed using the COG-UK tree generated on 25/02/2021. Estimates of growth rates of major lineages in Scotland were calculated from time-resolved phylogenies for lineages B.1.1.7 (Alpha), B.177 and the sub-clades B.177.5, B.177.8, and another minor B.177 sub-clade (W.4). The estimates were carried out utilising sequences from November 2020 – March 2021 in BEAST (Bayesian Evolutionary Analysis by Sampling Trees) with an exponential growth rate population model, strict molecular clock model and TN93 with four gamma rate distribution categories. Each lineage was randomly subsampled to a maximum of 5 sequences per epiweek (resulting in 52 to 103 sequences per subsample, depending on the lineage), and 10 subsamples replicates analysed per lineage in a joint exponential growth rate population model.

### Clinical data

we included all Scottish COG-UK pillar 1 samples sequenced at the MRC-University of Glasgow Centre for Virus Research (CVR) and the Royal Infirmary of Edinburgh (RIE) between 1st November 2020 and 30th January 2021. These samples derived from both hospitalised patients (59%) and community testing (41%).

Core demographic data (age, sex, partial postcode) were collected via linkage to electronic patient records and a full retrospective review of case notes was undertaken. Collected data included residence in a care home; occupation in care home or healthcare setting; admission to hospital; date of admission, discharge and/or death and maximum clinical severity at 28 days sample collection date via a 4-point ordinal scale (1. No respiratory support; 2. Supplemental oxygen; 3. Invasive ventilation, non-invasive ventilation or high-flow nasal canula; 4. Death) as previously used in Volz et al 2020 and Thomson et al 2021 (23,24).

Where available, PCR (Polymerase Chain Reaction) cycle threshold (Ct) and the PCR testing platform were recorded. Nosocomial COVID-19 was defined as a first positive PCR occurring greater than 48 hours following admission to hospital. Discharge status was followed up until 15th April 2021 for the hospital stay analysis. For the co-morbidity subanalysis, delegated research ethics approval was granted for linkage to National Health Service (NHS) patient data by the Local Privacy and Advisory Committee at NHS Greater Glasgow and Clyde. Cohorts and de-identified linked data were prepared by the West of Scotland Safe Haven at NHS Greater Glasgow and Clyde.

### Severity analyses

four level severity data was analysed using cumulative (per the definition of Bürkner and Vuorre (2019)) generalised additive mixed models (GAMMs) with logit links, specifically, following Volz et al (2020) (23,25). We analysed three subsets of the data: 1. the full dataset, 2. the dataset excluding care home patients, and 3. exclusively the hospitalised population. Further details regarding these analyses are provided in Supplementary Appendix 1.

### Ct analysis

Ct value was compared between Alpha variant and non-Alpha variant infections for those patients where the TaqPath assay (Applied Biosystems) was used. This platform was used exclusively for this analysis because different platforms output systematically different Ct values, and this was the most frequently used in our dataset (n = 154, Alpha = 38, non-Alpha = 116). We used a generalised additive model with a Gaussian error structure and identity link, and the same covariates used as in the severity analysis to model the Ct value. The model was fitted using the brms (v. 2.14.4) R package (26). The presented model had no divergent transitions and effective sample sizes of over 200 for all parameters. The intercept of the model was given a t-distribution (location = 20, scale = 10, df = 3) prior, the fixed effect coefficients were given normal (mean = 0, standard deviation = 5) priors, random effects and spline standard deviations were given exponential (mean = 5) priors.

### Hospital length of stay analysis

hospital length of stay was compared for Alpha variant and non-Alpha variant patients while controlling for age and sex using a Fine and Gray model competing risks regression using the crr function in the cmprsk (v. 2.2-10) R package (27,28). Nosocomial infections were excluded. In total, this analysis had 521 cases (Alpha = 187, non-Alpha = 334), of which 4 were censored; 352 patients were discharged from hospital and 165 died.

## Results

### Emergence of the Alpha variant in Scotland

Between 01/11/2020 and 31/01/2021 1863 samples from individuals tested in pillar 1 facilities in Scotland underwent whole genome sequencing for SARS-CoV-2. Of these, 1475 (79%) could be linked to patient records and were included in the analysis. The contribution of patients infected with the Alpha variant increased over the course of the study, in line with dissemination across the UK during the study period (Figure 1a and 1b). At the time of the data used in this analysis two peaks of SARS-CoV-2 infection had occurred in the UK: the first (wave 1) in March 2020 (15) and the second in summer 2020 (29), both in association with hundreds of importations following travel to Central Europe (30). The second peak incorporated two variant waves (waves 2 and 3), initially of B.1.177 (Figure 1c) and then B.1.1.7/Alpha, radiating from the South of England (Figure 1e). This Alpha variant “takeover” (Figure 1d) corresponded to a five-fold increase in growth rate on an epidemiological scale relative to non-Alpha lineages (Figure 1f).

**Figure 1.**
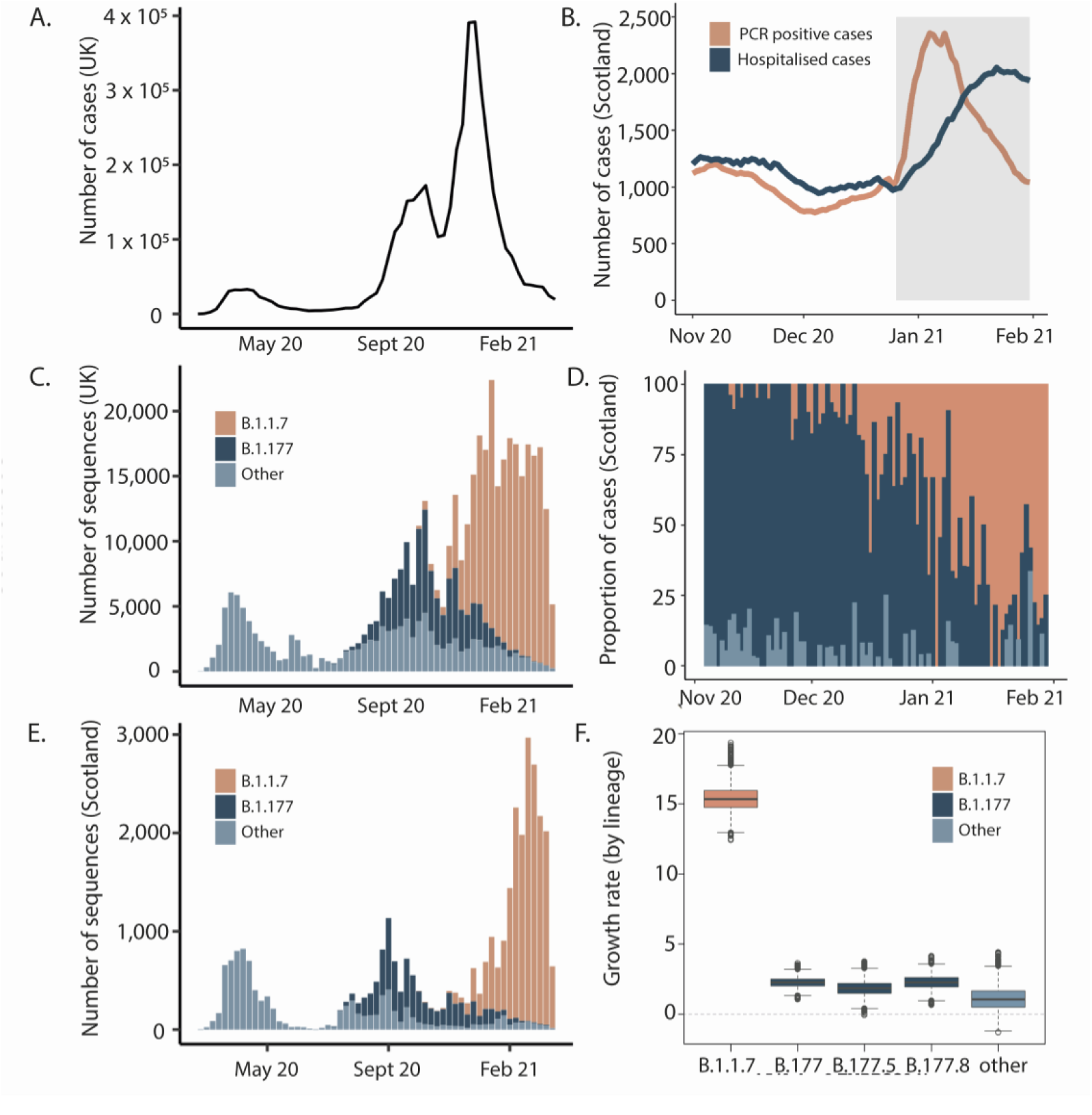
Introduction and growth of the Alpha variant (lineage B.1.1.7) in the UK. A) Waves of SARS-CoV-2 confirmed cases in the UK B) Seven-day rolling average of daily PCR positive cases (orange) and total number of patients hospitalised (dark blue) with COVID-19 in Scotland during the study period. Grey shaded area represents the period of lockdown beginning 26/12/2020 C) Variants in the UK D) Proportion of cases by lineage in the clinical severity cohort E) Variants in Scotland showing three distinct waves in winter and early spring 2020, summer 2020 and autumn/winter, attributed to the shifts from B1 and other variants (light blue) to B.1.177 (dark blue) and then B.1.1.7/Alpha (orange). Waves one and two closely mirror the broader UK situation as they are linked to both continental European and introductions from England. Wave three has a single origin in Kent so Scotland lags England in numbers of cases F) Estimates of growth rates of major lineages in Scotland from time-resolved phylogenies. Estimates were carried out on a subsample of the named lineages using sequences from Scotland only from November 2020-March 2021 using BEAST and an exponential growth effective population size model.

### Demographics of the clinical cohort

The age of the clinical cohort ranged from 0-105 years, (mean 66.8 years) and was slightly lower in the Alpha group (65.6 years vs. 67.2 years). Overall, 59.1% were female; this preponderance occurred in both subgroups and was higher in the Alpha subgroup (60.4% vs 58.6%). In the full cohort, 3.0% were care home workers and 10.4% were NHS healthcare workers. 5.5% and 5.8% of those infected with the Alpha variant were care home and other healthcare workers respectively, compared with 2.2% and 12.0% of those infected with non-Alpha lineages. 12.9% of those in the Alpha subgroup were care home residents, compared with 21.7% in non-Alpha. There was also a difference in the proportion of cases admitted to Intensive Care Units: 6.3% of the Alpha group compared with 3.4% for non-Alpha. Full details of the demographic data of the cohort can be found in Table 1 and full lineage assignments can be found in Table S2.

**Table 1:** Demographic characteristics of Scottish patients infected with SARS-CoV-2 by lineage.

| Characteristic | Overall Group<br>(n = 1475) |  | B.1.1.7 (Alpha)<br>(n = 364) |  | Other<br>(n = 1111) |  |
| --- | --- | --- | --- | --- | --- | --- |
|  | Number | Percentage | Number | Percentage | Number | Percentage |
| Age at diagnosis (years) |  |  |  |  |  |  |
| Mean ± SD | 66.8±20.8 |  | 65.6±20.6 |  | 67.2±20.8 |  |
| Range | 0-105 |  | 0-105 |  | 0-100 |  |
| Sex |  |  |  |  |  |  |
| Male | 604 | 40.9% | 144 | 39.6% | 460 | 41.4% |
| Female | 871 | 59.1% | 220 | 60.4% | 651 | 58.6% |
| Admitted to hospital |  |  |  |  |  |  |
| Yes | 876 | 59.4% | 238 | 65.4% | 638 | 57.4% |
| No | 599 | 40.6% | 126 | 34.6% | 473 | 42.6% |
| Care home worker |  |  |  |  |  |  |
| Yes | 44 | 3.0% | 20 | 5.5% | 24 | 2.2% |
| No | 1305 | 88.5% | 305 | 83.8% | 1000 | 90.0% |
| Unknown | 126 | 8.5% | 39 | 10.7% | 87 | 7.8% |
| Non-care home healthcare worker |  |  |  |  |  |  |
| Yes | 154 | 10.4% | 21 | 5.8% | 133 | 12.0% |
| No | 1193 | 80.9% | 305 | 83.8% | 888 | 79.9% |
| Unknown | 128 | 8.7% | 38 | 10.4% | 90 | 8.1% |
| Nursing home resident |  |  |  |  |  |  |
| Yes | 288 | 19.5% | 47 | 12.9% | 241 | 21.7% |
| No | 1187 | 80.5% | 317 | 87.1% | 870 | 78.3% |
| Unknown | 0 | 0.0% | 0 | 0.0% | 0 | 0.0% |
| Community transmission |  |  |  |  |  |  |
| Yes | 915 | 62.0% | 272 | 74.7% | 643 | 57.9% |
| No | 376 | 25.5% | 44 | 12.1% | 332 | 29.9% |
| Unknown | 184 | 12.5% | 48 | 13.2% | 136 | 12.2% |
| Diagnosis >48 hours post-admission |  |  |  |  |  |  |
| Yes | 346 | 23.5% | 46 | 12.6% | 300 | 27.0% |
| No | 1040 | 70.5% | 289 | 79.4% | 751 | 67.6% |
| Unknown | 89 | 6.0% | 29 | 8.0% | 60 | 5.4% |
| Travel outside Scotland |  |  |  |  |  |  |
| Yes | 1 | 0.1% | 0 | 0.0% | 1 | 0.1% |
| No | 317 | 21.5% | 20 | 5.5% | 297 | 26.7% |
| Unknown | 1157 | 78.4% | 302 | 94.5% | 813 | 73.2% |
| Immunosuppressed |  |  |  |  |  |  |
| Yes | 42 | 2.9% | 4 | 1.1% | 38 | 3.4% |
| No | 474 | 31.1% | 60 | 16.5% | 414 | 37.3% |
| Unknown | 959 | 65.0% | 300 | 82.4% | 659 | 59.3% |
| Visited Intensive Care Unit? |  |  |  |  |  |  |
| Yes | 61 | 4.1% | 23 | 6.3% | 38 | 3.4% |
| No | 1413 | 95.8% | 341 | 93.7% | 1072 | 96.5% |
| Unknown | 1 | 0.1% | 0 | 0.0% | 1 | 0.1% |
| Patient alive/deceased? |  |  |  |  |  |  |
| Alive | 1115 | 75.6% | 273 | 75.0% | 842 | 75.8% |
| Deceased | 360 | 24.4% | 91 | 25.0% | 269 | 24.2% |

### Clinical severity analysis

Within the clinical severity cohort there were 364 Alpha, 1030 B.1.177 and 81 of 19 other lineage infections (Figure 2). Consistent with previous research comparing mortality and hospitalisation in SGTF detected by PCR versus absence of SGTF, we found that Alpha variant viruses were associated with more severe disease on average than those from other lineages circulating during the same time period. In the full dataset, we observed a positive association with severity (posterior median cumulative odds ratio: 1.40, 95% CI: 1.02-1.93). In both the subsets excluding care home patients, or limiting to hospitalised patients, the mean estimate of the increase in severity of the Alpha variant was smaller, and the variance in the posterior distribution higher likely due to the smaller sample sizes. Given this uncertainty, we cannot determine whether the association of the Alpha variant with severity in the populations corresponding to these subsets is the same as that in the population described by the full dataset, but in all cases, the most likely direction of the effect is positive. Model estimates from severity models from all subsets can be found in Tables S3-5.

**Figure 2:**
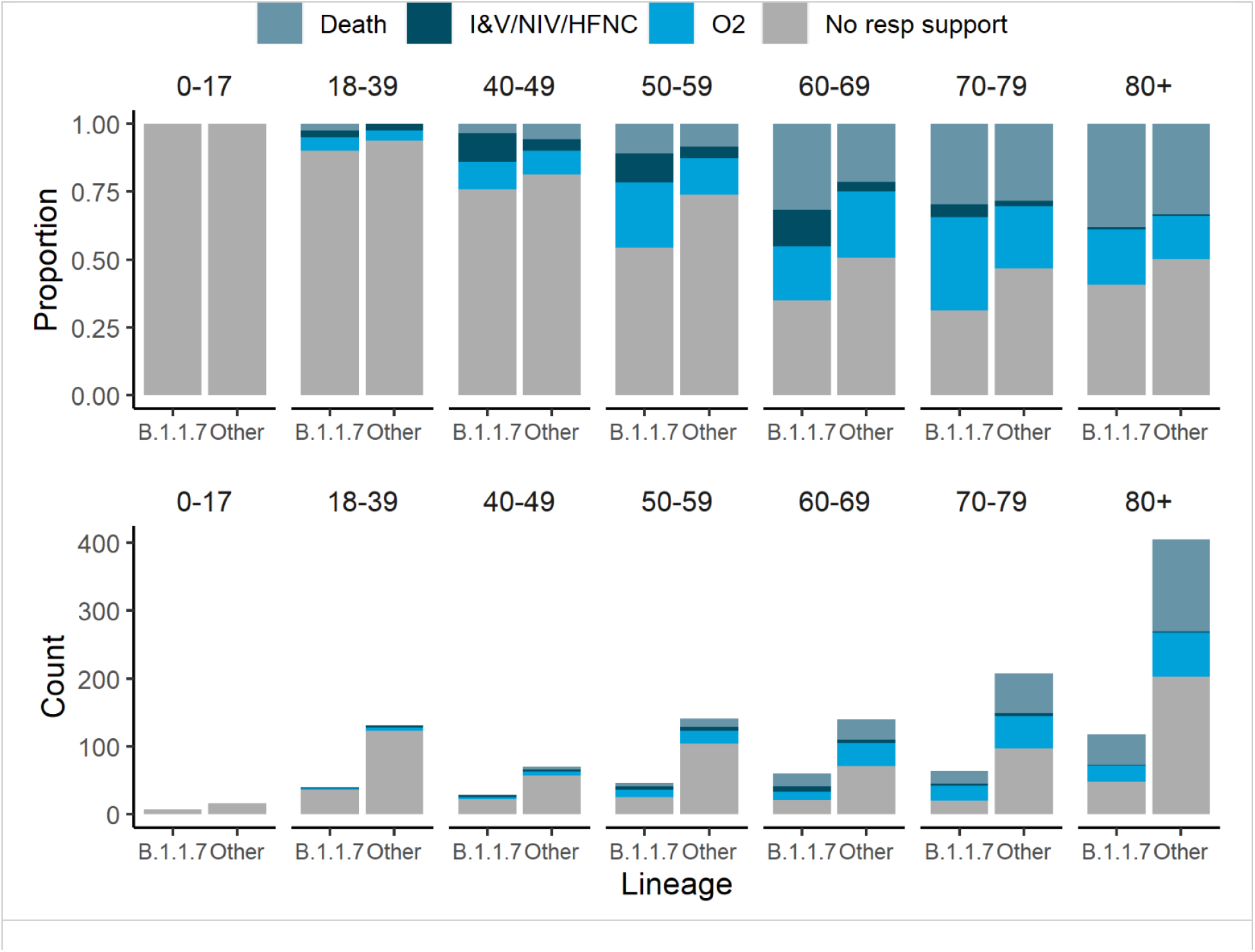
Comparison of disease severity between the Alpha variant (B.1.1.7) and other lineages. Clinical severity was measured on a four-level ordinal scale based on the level of respiratory support received for 1454 patients stratified by age group; death, invasive or non-invasive ventilatory support including high flow nasal cannulae (I&V/NIV/HFNC), supplemental oxygen delivered by low flow mask devices or nasal cannulae, and no respiratory support.

Bernoulli models looking at sequential severity categories provided weak evidence that the proportional odds assumption of the cumulative logistic model was violated. The odds ratios for the no oxygen versus low flow oxygen, and low flow oxygen versus were similar to those estimated under the cumulative model (posterior median odds ratio for no oxygen versus low flow oxygen: 1.77, CI: 1.12-2.80; posterior median odds ratio for low flow oxygen versus high flow oxygen: 1.26, CI: 0.43-3.67) but with correspondingly higher posterior variances given the smaller sample size. The odds ratios for the high flow oxygen versus death model suggested that the preponderance of evidence was in favour of Alpha infection associated with lower risk of death, conditional on having received high flow oxygen (posterior median odds ratio: 0.64, CI: 0.22-1.90). However, the credible intervals here are wide, given the sample size and do include the estimated global effect. A similar but more extreme effect was observed for the effect of biological sex, with male sex being associated with worse outcomes for the first two sequential category models (posterior median odds ratio for no oxygen versus low flow oxygen: 1.32, CI: 0.96-1.80; posterior median odds ratio for low flow oxygen versus high flow oxygen: 3.10, CI: 1.37-7.08), but better outcomes for the last (posterior odds ratio: 0.62, CI: 0.19-099). Given other research on the topic has consistently identified male sex as a risk factor, this potentially indicates the existence of an important unmeasured confounder only relevant for those who receive high flow oxygen.

Estimates of the severity across the phylogeny are visible in Figure 3, see Supplementary Appendix 2 for more discussion of this analysis. An analysis including comorbidities for the subset of patients where they were available implied that the inclusion of comorbidities had no impact on the results obtained, see Supplementary Appendices 1 and 3.

**Figure 3:**
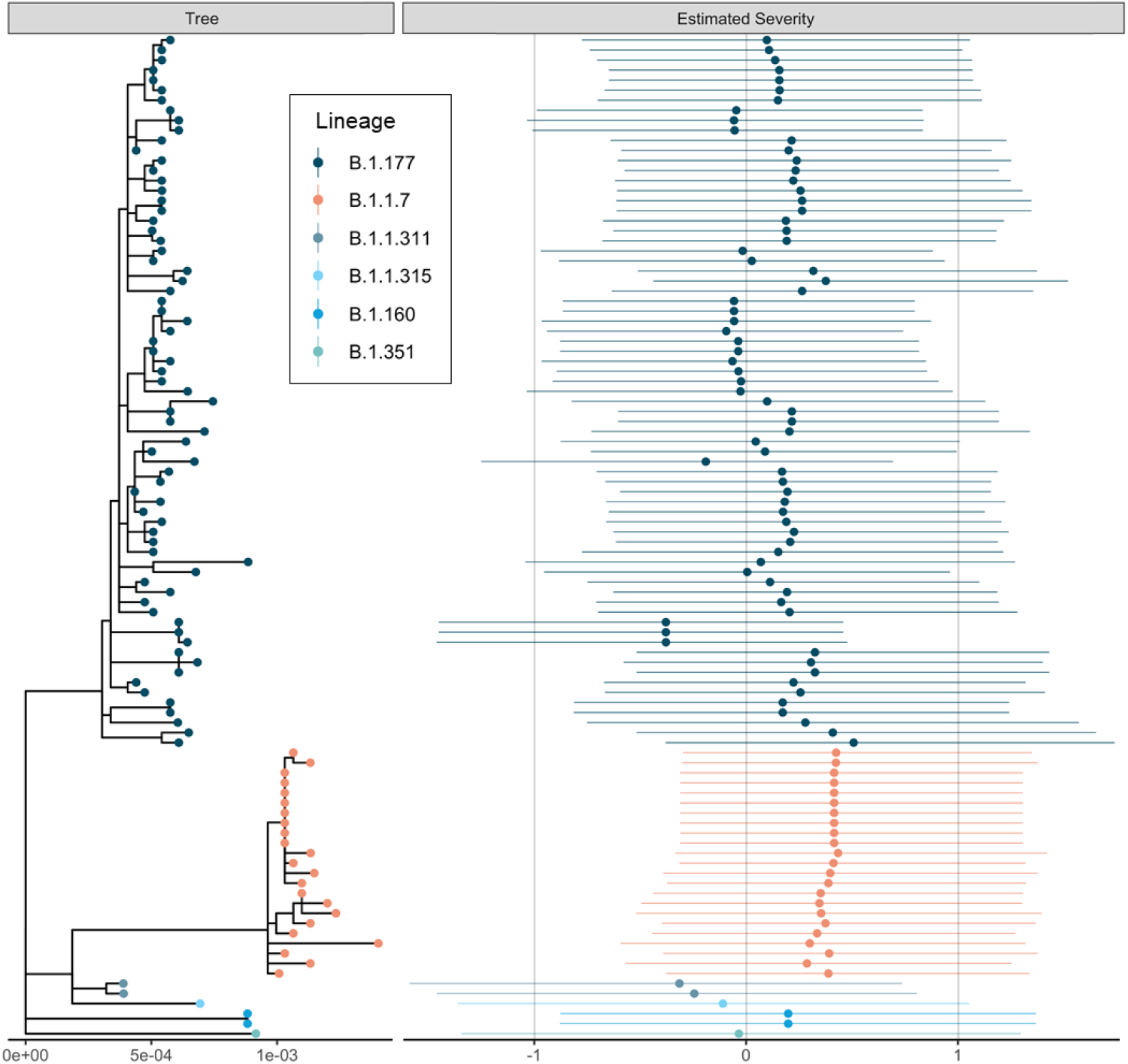
The estimated maximum likelihood phylogenetic tree and a measure of estimated severities of infection. Estimated severities for each viral isolate are means and 95% credible intervals of the linear predictor change under infection with that viral genotype from the phylogenetic random effect in the cumulative severity model under a Brownian motion model of evolution. This model constrains genetically identical isolates to have identical effects, so changes should be interpreted across the phylogeny rather than between closely related isolates which necessarily have similar estimated severities. The dataset was downsampled to 100 random samples for this figure to aid readability. Figure was generated using ggtree (31).

We also found that the Alpha variant was associated with lower Ct values than infection with non-Alpha variants (posterior median Ct change: -2.46, 95% CI: (−4.22)-(−0.70)) as previously observed (8). Model estimates for all parameters can be found in Table S6.

We found no evidence that the Alpha variant was associated with longer hospital stays after controlling for age and sex (HR: -0.02; 95% CI: (−0.23)-0.20; p = 0.89).

## Discussion

In this analysis of hospitalised and community patients with Alpha variant and non-Alpha variant SARS-CoV-2 infection, carried out as the Alpha variant became dominant in Scotland, we provide evidence of increased clinical severity associated with this variant. This was observed across all adult age groups, incorporating the spectrum of COVID-19 disease; from no requirement for supportive care to supplemental oxygen requirement, the need for invasive or non-invasive ventilation, and to death. This analysis is the first to assess the full clinical severity spectrum of confirmed Alpha variant infection in both community and hospitalised cases in relation to other prevalent lineages circulating during the same time period.

Our study supports the community testing analyses that have reported an increased 28-day mortality associated with SGTF as a proxy for Alpha variant status (8-10). Smaller studies found no effect of lineage on various measures of severity (15-17), but these were studies of patients already admitted to hospital and therefore would not pick up the granular detail of increasing disease severity resulting in a need for increasing levels of respiratory support and consequently admission to hospital.

The association between higher viral load, higher transmission and lineage may reflect changes in the biology of the virus; for example, the Alpha variant asparagine (N) to tyrosine (Y) mutation at position 501 of the spike protein receptor binding domain (RBD) is associated with an increase in binding affinity to the human ACE2 receptor (32). In addition, a deletion at position 69–70 may increase virus infectivity (33). The P681H mutation found at the furin cleavage site is associated with more efficient furin cleavage, enhancing cell entry (34). An alternative explanation for the higher viral loads observed in Alpha variant infection may be that clinical presentation occurs earlier in the illness. Further modelling, animal experiments and studies in healthy volunteers may help to unravel the mechanisms behind this phenomenon.

Our data indicate an association between the Alpha variant and an increased risk of requiring supplemental oxygen and ventilation; two factors that are critical determinants of healthcare capacity during a period of high incidence of SARS-CoV-2 infection. This illustrates the importance of countries, in particular those with weaker public health control of the virus, factoring the requirement for supportive treatment into models of clinical severity and pandemic response decision planning for future SARS-CoV-2 variants of concern. This granular analysis of disease severity based on genomic confirmation of diagnosis should be used as a baseline study for clinical severity analysis of the inevitable future variants of concern.

There are some limitations to our study. Our dataset is drawn from first-line local NHS diagnostic (Pillar 1) testing which over-represents patients presenting for hospital care (59%) while those sampled in the community represented 41% of the dataset. Further, the analysis dataset employed a non-standardised approach to sampling across the study period as sequencing was carried out both as systematic randomised national surveillance and sampling following outbreaks of interest. Finally, the cumulative model used in this analysis assumes a homogenous application of therapeutic intervention across the population. Despite these limitations, our results remain consistent with previous work on the mortality of Alpha, and this study provides new information regarding differences in infection severity.

In summary, the Alpha variant was found to be associated with a rapid increase in SARS-CoV-2 cases in Scotland and an increased risk of severe infection requiring supportive care. This has implications for planning for future variant driven waves of infection, especially in countries with low vaccine uptake or if variants evolve with significant vaccine-escape. Our study has shown the value of the collection of higher resolution patient outcome data linked to genetic sequences when looking for clinically relevant differences between viral variants.

## Data Availability

Due to the analysis of patient identifiable data, please contact the authors for data requests.

## Acknowledgements

We would like to thank all NHS staff that looked after patients during the COVID-19 pandemic in Scotland. The authors would like to acknowledge that this work uses data provided by patients and collected by the National Health Service (NHS) as part of their care and support. The authors would also like to acknowledge the work of the West of Scotland Safe Haven team in supporting extractions and linkage to de-identified NHS patient datasets.

### Supplementary Appendix

**Table S1:** Characteristic mutations of the Alpha variant.

| Gene | Amino acid |
| --- | --- |
| ORF1a | T1001I |
| ORF1a | A1708D |
| ORF1a | I2230T |
| ORF1a | del3675/3677 |
| ORF1b | P314L |
| S | del69/70 |
| S | del144/145 |
| S | N501Y |
| S | A570D |
| S | D614G |
| S | P681H |
| S | T716I |
| S | S982A |
| S | D1118H |
| ORF8 | Q27* |
| ORF8 | R52I |
| ORF8 | Y73C |
| N | D3L |
| N | R203K |
| N | G204R |
| N | S235F |

**Table S2:** Full lineage characterisation of clinical severity dataset.

| Lineage | Count |
| --- | --- |
| A.23.1 | 1 |
| B.1 | 1 |
| B.1.1.1 | 4 |
| B.1.1.10 | 1 |
| B.1.1.163 | 1 |
| B.1.1.250 | 1 |
| B.1.1.311 | 18 |
| B.1.1.315 | 6 |
| B.1.1.37 | 11 |
| B.1.1.7 (Alpha) | 364 |
| B.1.160 | 13 |
| B.1.177 | 1030 |
| B.1.2 | 1 |
| B.1.221 | 2 |
| B.1.235 | 2 |
| B.1.258 | 10 |
| B.1.351 | 3 |
| B.1.36 | 3 |
| B.1.389 | 1 |
| B.1.88 | 1 |
| P.2 | 1 |

**Table S3:** Parameter estimates (on the linear predictor scale) from the severity model from the full dataset.

|  | Median | Lower Bound | Upper Bound |
| --- | --- | --- | --- |
| Intercept 1 | 0.81 | 0.55 | 1.10 |
| Intercept 2 | 1.7 | 1.44 | 2.01 |
| Intercept 3 | 1.89 | 1.62 | 2.21 |
| Alpha variant | 0.34 | 0.02 | 0.66 |
| Male Sex | 0.45 | 0.23 | 0.68 |
| Linear effect of age | 1.18 | -0.34 | 3.28 |
| Linear effect of date | -0.09 | -0.81 | 0.14 |

**Table S4:** Parameter estimates (on the linear predictor scale) from the severity model from the data subset excluding patients in nursing homes.

|  | Median | Lower Bound | Upper Bound |
| --- | --- | --- | --- |
| Intercept 1 | 0.86 | 0.52 | 1.28 |
| Intercept 2 | 1.89 | 1.55 | 2.33 |
| Intercept 3 | 2.14 | 1.79 | 2.58 |
| Alpha variant | 0.17 | -0.21 | 0.55 |
| Male Sex | 0.50 | 0.26 | 0.75 |
| Linear effect of age | 1.16 | -0.52 | 3.40 |
| Linear effect of date | -0.03 | -0.48 | 0.08 |

**Table S5:**
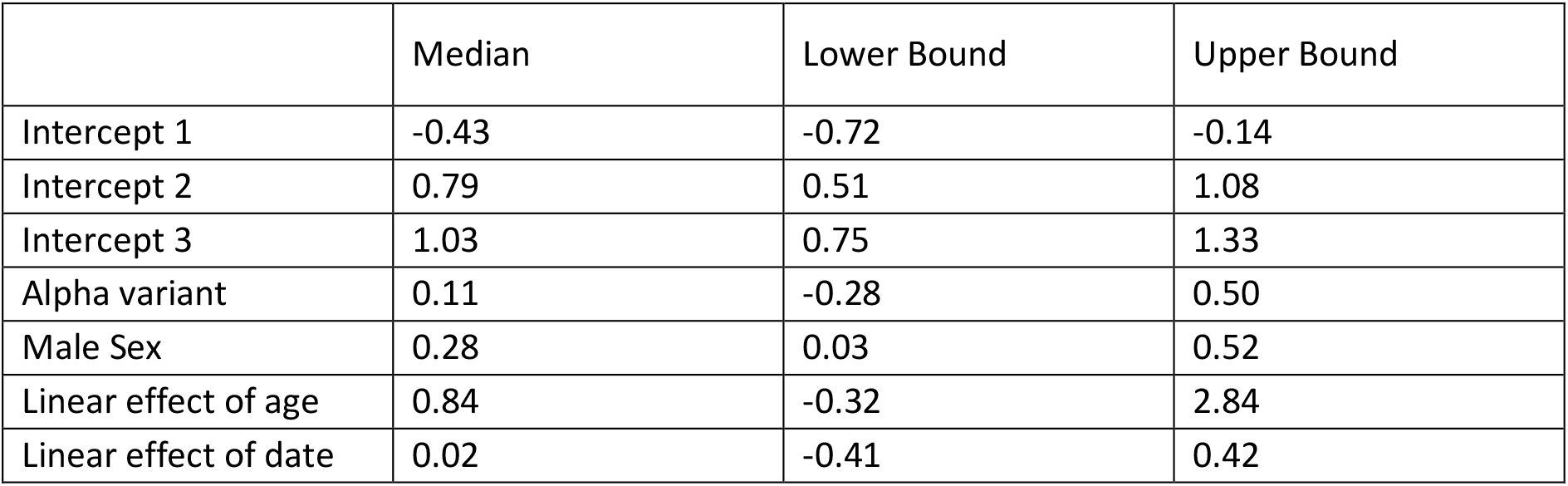
Parameter estimates (on the linear predictor scale) from the severity model from the data subset only including hospitalised patients.

**Table S6:** Parameter estimates from the Ct value model.

|  | Median | Lower Bound | Upper Bound |
| --- | --- | --- | --- |
| Intercept | 21.95 | 16.69 | 23.66 |
| Alpha variant | -2.46 | -4.22 | -0.70 |
| Male Sex | 0.70 | -0.78 | 2.18 |
| Linear effect of age | 0.02 | -0.56 | 1.09 |
| Linear effect of date | 0.12 | -0.57 | 1.23 |

### Appendix 1 – Further methods

Following Volz et al (2020), we modelled the ordinal severity data using cumulative generalised additive mixed models (per the definition of (per the definition of Bürkner and Vuorre (2019)) (18,20). We analysed three subsets of the data: 1. the full dataset, 2. the dataset excluding care home patients, and 3. exclusively the hospitalised population. The impact of Alpha variant infection and patient sex were modelled with fixed effects. County and partial postcode were modelled as random effects. Patient age and the days since the first diagnosis in the dataset were modelled using non-linear penalised regression splines with the *k* parameter set to its maximum value. The full dataset was additionally analysed using a phylogenetic cumulative generalised additive mixed model (PGAMM). The PGAMM was a modification of the GAMMs described above, where instead of including Alpha variant status as a fixed effect, we included a random effect of phylogenetic relationship between viral isolates (using a variance-covariance matrix calculated from the virus phylogeny under a Brownian motion assumption using the vcv.phylo function in ape (v. 5.5) (1)). All severity models were fitted using the brms (v. 2.14.4) R package (2). All presented models had no divergent transitions and effective sample sizes of over 200 for all parameters. Additionally, we fitted Bernoulli models with the same covariate set as the cumulative model for no oxygen vs. low flow supplemental oxygen, low flow supplemental oxygen vs. high flow supplemental oxygen and, high flow supplemental oxygen vs. mortality individually to test the proportional odds assumption.

Comorbidities were only available for patients from the Greater Glasgow and Clyde health board (n = 639). Comorbidities used were those previously identified as important for COVID-19 severity by the ISARIC4C consortium (3). To test whether the lack of comorbidity data for the rest of the sample was leading to biased estimates of the impact of Alpha variant infection, we performed three analyses on the Greater Glasgow and Clyde patient population. We fit the above model with the number of comorbidities a patient exhibited included as non-linear penalised regression spline. While the exact form of the relationship between severity of infection and the number of comorbidities a patient exhibits is unknown, we would expect the relationship to be monotonically increasing, however, for mathematical simplicity, we do not enforce this constraint on the spline. We also fit the model to this patient population without the comorbidities included and with the comorbidities permuted to estimate the change in the estimate of the Alpha variant effect by the inclusion of comorbidities. As the inclusion of comorbidities was found not to change the estimated effect of the Alpha variant, this analysis is presented in Supplementary Appendix 4.

Model intercepts were given t-distribution (location = 0, scale = 2.5, df = 3) priors, fixed effects were given normal (mean = 0, standard deviation = 2.5) priors, random effects and spline standard deviations were given exponential (mean = 2.5) priors.

## Appendix 2 – Phylogenetic severity model

The estimates of the severity per isolate shown in Figure 3 were generated by a model making several assumptions, which were violated. The key assumptions used, and their impacts will be discussed in this appendix (see 1 for deeper discussion of some the issues involved). Despite the violation of the assumptions, the answer generated was consistent with the non-phylogenetic method and the output is illustrative, so the results are included in the main text, though not stressed.

The first major assumption is that the source phylogeny is known without error. This can be practically broken into two assumptions. Firstly, that tree-like evolution is the correct description of the underlying evolutionary process, i.e., that horizonal gene transfer is unimportant. This is a relatively safe assumption in SARS-CoV-2. Secondly, that the phylogenetic tree is correctly estimated. This is likely to be violated as there may be error in both the discrete branching structure (or topology) and real-valued branch lengths. While the topology may be correctly estimated, the probability of estimating all the branch lengths correctly is vanishingly small. This is unlikely to be a large practical issue however, as small errors in the branch lengths of the phylogeny are unlikely to have large impacts relative to other model misspecification issues present in all statistical analyses.

If we are willing to assume that the estimated phylogeny is good enough for our purposes, we then must assume some model of the evolution of the trait of interest across that phylogeny. This model of the change in the trait (in this case, severity) across the phylogeny is what allows the conversion of the phylogenetic tree into a variance-covariance matrix. This describes the expected covariances (rescaled to correlations) between the severities associated with infection with different genetic variants. Here we made a common simple choice and assumed Brownian motion evolution of the trait across the phylogeny. However, this model has been acknowledged as often suboptimal since its inception (1), and we can consider it particularly so here. The number of observed changes across SARS-CoV-2 genomes are relatively few, and the number of amino acid changes even fewer, with some mutations occurring repeatedly in different lineages. Few mutations with combined with semi-frequent homoplasy represent a particularly problematic case for this model, as severity would be expected to change discretely with mutations and in consistent directions when convergent changes occur (in the absence of extreme epistatic effects on severity), two things that simple Brownian motion does not allow. Future work will explore more realistic evolutionary models for change in severity with genomes, such as matching criteria, which will reduce the error potentially imposed by this assumption.

## Appendix 3 - Comorbidities

In the Greater Glasgow and Clyde population for which comorbidity data was available, the model without inclusion of comorbidities estimated the odds ratio for the impact of the Alpha variant on severity as 1.06 (95% CI: 0.70, 1.58). When number of relevant comorbidities a patient had were included but permuted, to break any relationship with the response, a similar odds ratio was estimated (1.06: 95% CI: 0.70, 1.60). The inclusion of the number of relevant comorbidities a patient exhibited did not substantially change this result (odds ratio for impact of the Alpha variant: 1.13; 95% CI: 0.73, 1.72). This is not unexpected, as the distribution of comorbidities was similar between those patients infected with the Alpha variant and those infected with non-Alpha lineage viruses.

